# Rationale and design of the Cancer Immunotherapy Evidence Living (CIEL) Library: A continuously updated clinical trial database of cancer immunotherapies

**DOI:** 10.1101/2024.04.26.24306436

**Authors:** K Boesen, J Hirt, P Düblin, H Läubli, B Kassenda, LG Hemkens, P Janiaud

## Abstract

**Background:** Immunotherapies for cancers are being tested in large numbers of clinical trials. It is nearly impossible for clinicians and researchers to stay current with the evidence, and traditional systematic reviews and clinical guidelines are not suited to ensure a continued overview of all trials and their results. To address this problem, we have designed a free-to-use, and publicly available database of clinical trials that aims to be continuously updated, the Cancer Immunotherapy Evidence Living (CIEL) Library.

**Methods:** We aimed to include planned, ongoing, and completed interventional trials of immunotherapies for cancer, regardless of trial design (e.g., randomization, blinding, and type of comparator). We systematically searched PubMed (for published reports) and ClinicalTrials.gov (for registered clinical trials). PubMed retrieved records were screened using the AI-assisted software ASReview and manually extracted and curated. We imported data from ClinicalTrials.gov using the Clinical Trials Transformation Initiative database which then requires further curation. The CIEL-Library is available and searchable via a web application (https://app.ciel-library.org/#/). It also contains the ‘Match My Patient’ feature, a patient-centered clinical decision support system, which can filter planned, ongoing or completed trials based on four patient characteristics (disease staging, previous treatments, performance status, and location). We piloted our database with one type of cancer immunotherapy, the tumor-infiltrating lymphocytes (TIL) transfer.

**Conclusion:** The CIEL-Library offers a blueprint for a dynamic evidence synthesis infrastructure providing an exhaustive collection of clinical trials with trial characteristics and results, which can be applied across different fields, specialties, and topics. The Match My Patient search function may be very useful to implement trial research into patient-centered care by helping to find a trial for enrolment of a patient or to find results for making decisions, for example, in tumor boards.

The main challenges to making a continuously updated database of clinical trials are the time and resources needed to populate it with curated and updated data. The CIEL-Library project illustrates the potential and the main limitations to designing such continuously updated trial databases that intent to be directly used in routine care.

## Background

Treatment options for cancer patients have substantially increased, especially in the field of immunotherapies. Immunotherapies for cancer include small molecule agents (e.g. IDO (indoleamine 2,3-dioxygenase) inhibitors), checkpoint inhibitors (e.g. PD-1 and CTLA4 inhibitors), and adoptive cell therapies (ACT; e.g. tumor-infiltrating lymphocytes (TIL), chimeric antigen receptor (CAR) T cells and engineered T cells, and therapeutic cancer vaccines, (e.g. dendritic and DNA/mRNA-based vaccines)) (1–4).

The number of planned, ongoing, and completed clinical trials investigating immunotherapy has increased drastically. In 2018, 1388 new trials were initiated across all types of immunotherapies for cancer, and in 2022 this number rose to 2095 new trials (5,6). The most prolific immunotherapeutic option is CAR T-cells with, in December 2020, 778 trials registered on ClinicalTrials.gov (7). The US Food and Drug Administration (FDA) approved six CAR-T cell products for different lymphoma, multiple myeloma, and leukemia indications between 2017 and 2022 (8). Three therapeutic cancer vaccines have also received FDA approval; the Bacillus Calmette-Guérin vaccine for bladder cancer in 1990, a dendritic cell-based vaccine (sipuleucel-T) for prostate cancer in 2010 (9), and an oncolytic herpes simplex virus (talimogene laherparepvec) for melanoma in 2022 (10).

Immunotherapies for cancer is a rapidly evolving field with an escalating literature. Thus, it is time consuming and laborious for individual researchers to get a comprehensive overview and to evaluate key trial characteristics. Similarly in clinical settings, complex oncology patient cases are often discussed at interdisciplinary tumor board meetings to decide on the most evidence-based and patient-tailored treatment. Such clinical decisions face the same challenges to identify the most relevant evidence. Tools that support research and clinical needs to identify and appraise the most current available evidence efficiently, reliably, and transparently are needed.

Treatment decision-making builds on clinical expertise, patient preferences, and available evidence (11). Databases of published research, such as PubMed, and clinical trial registries, such as ClinicalTrials.gov, have improved access to information about published and planned clinical trials. However, the number of clinical trials testing healthcare interventions increases exponentially and it is impossible for clinicians to stay on top of the evidence (12). Clinical guidelines and systematic reviews also cannot reflect the rapid pace of evidence generation. Systematic reviews may be outdated even before publication (13) or not kept up-to-date, (14) likely because it is time and resource demanding to manually search, screen, assess, and extract data from new records.

An increasing number of tools and software exist to automatize and streamline literature screening and data extraction in the systematic review process (15,16), but these tools largely root in the systematic review being a ‘static’ publication. In contrast to such static reviews, one potential solution is to design a continuously updated database of clinical trials, automatizing as many steps as possible.

### The Cancer Immunotherapy Evidence Living (CIEL) Library

We aimed to develop a web application for a comprehensive, continuously updated living library of all related immunotherapy clinical trials (planned, ongoing and completed) and corresponding results to guide clinical decisions and clinical research, and to make it freely accessible. We planned to (semi)automatize as many steps as possible to reduce the burden of maintenance.

The CIEL-Library should serve a triple purpose as (i) an exhaustive library of clinical trials; (ii) a research resource to highlight gaps in the clinical research agenda and evidence base; and (iii) a clinical decision support system, (17–19) with potential direct implementation in the clinic. It has two main features the ***CIEL-Library*** and ***Match My Patient***.

The ***CIEL-Library*** is the actual library, which contains all identified trials assessing our immunotherapies of interests. The library is searchable, and the trials can be filtered. For each trial, we present general characteristics and key details following the PICO framework (i.e., Population, Intervention, Comparator and Outcome) and a direct link to the original record. All data underlying the CIEL-Library is available in a structured format, ready to be used for research purposes, e.g. meta-epidemiological assessments of immunotherapy clinical trials. This may enable identification of research gaps and highlight strengths and limitations of the research literature.

The ***Match My Patient*** search function is intended as a patient-centered clinical decision support system to help identify ongoing (i.e., to find a trial to enroll yourself or your patient) or completed (i.e., to find results for making a therapeutic decision) clinical trials based on patient characteristics. A clinical decision support system can be defined as a platform/software where patients with specific characteristics, e.g. age, sex, biomarkers, and disease severity, are compared with a ‘knowledge base’ to guide decision making (17). Such knowledge base can be a clinical guideline, records of previously treated patients, or in our case, an exhaustive collection of clinical trials. See use case example in Box 1.

#### BOX 1.

Clinical use case of CIEL’s *Match my patient* feature.

An oncologist wants to find clinical evidence on the potential use of tumor-infiltrating lymphocytes for a patient with advanced melanoma (TNM stage IV), who has already received one line of systemic treatment, and is fully active (performance status 0). Using *Match My Patient*, the oncologist can identify trials which match the patient characteristics (See Figure 4 for an example). At the subsequent tumor board meeting, the oncologist presents the available evidence using the CIEL-Library web application live.

## Methods

In this rational and design paper, we highlight the CIEL-Library’s overall infrastructure with its four main workstreams: (i) Retrieval and screening of PubMed records, (ii) retrieval and screening of ClinicalTrials.gov records, (iii) designing the CIEL database, and (iv) designing the CIEL-Library (Figure 1). We leveraged and optimized the implementation and methodology of previously established databases also led by us on COVID-19 trials (20), pragmatic trials (21,22), and on FDA regulatory documents pertaining to cancer therapies approvals (23,24).

**Figure 1:**
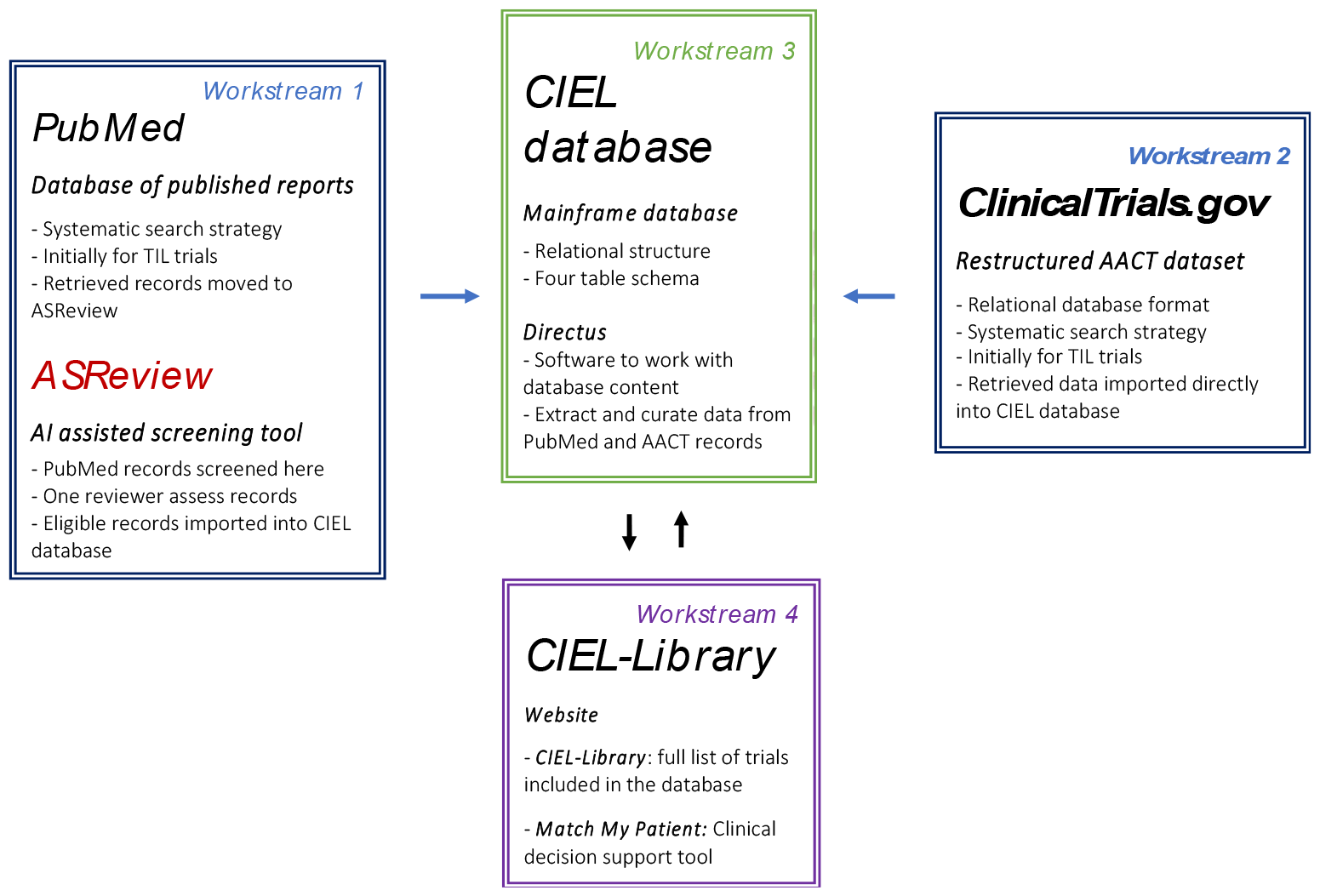
CIEL-Library workstreams

### Eligibility criteria

The CIEL-Library aims to contain any intervention study (trial) regardless of study phase, number of arms, randomization or not, blinding or not, and study design, assessing an immunotherapeutic intervention against any comparator (i.e. placebo, active drug, usual care, or no control) for any hematological or solid cancer. We designed and piloted the database with one specific type of immunotherapy, tumor-infiltrating lymphocytes (TIL) transfer (25,26). TILs are immune cells extracted from patient tumor tissue and then expanded (grown) in-vitro under specific circumstances for weeks before being injected into the patients again. TIL therapy has mainly been tested in metastatic melanoma patients (27) and non-small cell lung cancer patients (28). We chose this therapeutic option due to the anticipated number of trials (not too many, not too few) (29).

### Data sources and search strategy

Our strategy was to implement a systematic search approach for each data source following best methodological practices from systematic reviews. We focused on PubMed as the source for published trial reports of results and ClinicalTrials.gov as the source for registered planned, ongoing, and completed trials. We assumed that these sources cover the most impactful research studies in the field.

We (JH, PJ, and KB) systematically designed and verified a search string with a pre-emptive list of keywords for the condition (‘any cancer disease’) and intervention (‘tumor-infiltrating lymphocytes’), see Appendix 1. Search terms were collected by expert consultation (BK) and initial searches on the topic using PubMed, Epistemonikos, PROSPERO, Scopus, and Google Scholar. In addition, we identified controlled vocabulary using the MeSH-browser. PubReMiner (30) was also used to identify additional free text terms and controlled vocabulary that has been used in relevant publications in the field.

### Identification and screening of PubMed records (workstream 1)

To reduce the time needed to screen and identify relevant trials, we implemented a machine-learning assisted screening tool, ASReview (31,32). It is an open-source tool developed at Utrecht University, the Netherlands. It uses a machine-learning algorithm to rank the references obtained from a database search according to their relevance. Instead of having to go through all references randomly, ASReview presents the most relevant reference first, and the least relevant reference last.

For our initial search for tumor-infiltrating lymphocyte trials, we searched PubMed and retrieved 14,004 records (2 May 2023). We employed a two-step screening process. We first restricted our total sample adding the PubMed filter “Clinical study” to our search and screened all 604 resulting records. We subsequently used the trained ASReview’s algorithm on those 604 records to screen the full PubMed sample. We had predefined an arbitrary stopping rule of 100 consecutive non-eligible records. Ultimately, we screened a total of 1,476 records (11% of the total sample) and we identified 136 relevant records (0,97% of the total sample).

### Identification and screening of ClinicalTrials.gov records (workstream 2)

ClinicalTrials.gov is the single-most comprehensive trial registry with more than 480,000 trials registered by January 2023. For data import, we used the Clinical Trials Transformation Initiative’s database (AACT), which is an up-to-date copy of all information on ClinicalTrials.gov made available in a relational database format (33,34). Compared to using ClinicalTrials.gov’s native application programming interface (API) to import data directly into our CIEL database, the AACT’s relational database format (in theory) allows a more comprehensive and smooth integration, in addition to also containing more data.

Practically, we downloaded the AACT dataset as a static database dump on a monthly basis, which consists of 47 data tables in text file (.txt) format. The data dump is updated daily on AACT’s website, and AACT also makes available a cloud-based version of the database, which is also updated daily. We searched the tables “interventions.txt” and “interventions_other_names.txt.” using the same keywords derived from our PubMed literature search string (Appendix 1). Our most recent systematic search (31 January 2024) returned 270 trials. Retrieved records are automatically imported into the CIEL database and are then checked for eligibility during data curation. Overall, 179 trials (66 %) were eligible and 91 hits (34%) were not eligible, e.g. not an interventional trial or not the right population, e.g. treatment for psoriasis.

### CIEL Database architecture (workstream 3)

We designed the underlying CIEL Database as a relational database. A relational database combines multiple tables (or spreadsheets) and allows cross-referencing across these tables through unique identifiers (35). In contrast to single-sheet non-relational tables (like Excel spreadsheets), this structure also allows to continuously update and increase the database content.

Our software engineer (PD) built a relational Structured Query Language database using PostgreSQL as the relational database management system, which is an open-source non-proprietary program (PostgreSQL). We made a five-table database schema (Figure 2): Table 1 (‘Trial information’; 52 variables; 29 automatically imported from AACT) contains all basic trial information; Table 2 (‘Groups’; 19 variables; 4 automatically imported) contains all information related to study groups, e.g. allocated treatments and also baseline information (provided it has been reported on a group level); Table 3 (‘Outcomes’; 7 variables; 4 automatically imported) describes the assessed and reported trial outcomes; Table 4 (‘Group results’; 7 variables; 5 automatically imported) contains results at the study group level; and Table 5 (‘Results’; 9 variables; 6 automatically imported) contains the corresponding comparison results. The codebook underlying the database is available from the Open Science Network (36), in which all 82 variables are specified, also those variables that currently do not directly appear on the web application.

**Figure 2:**
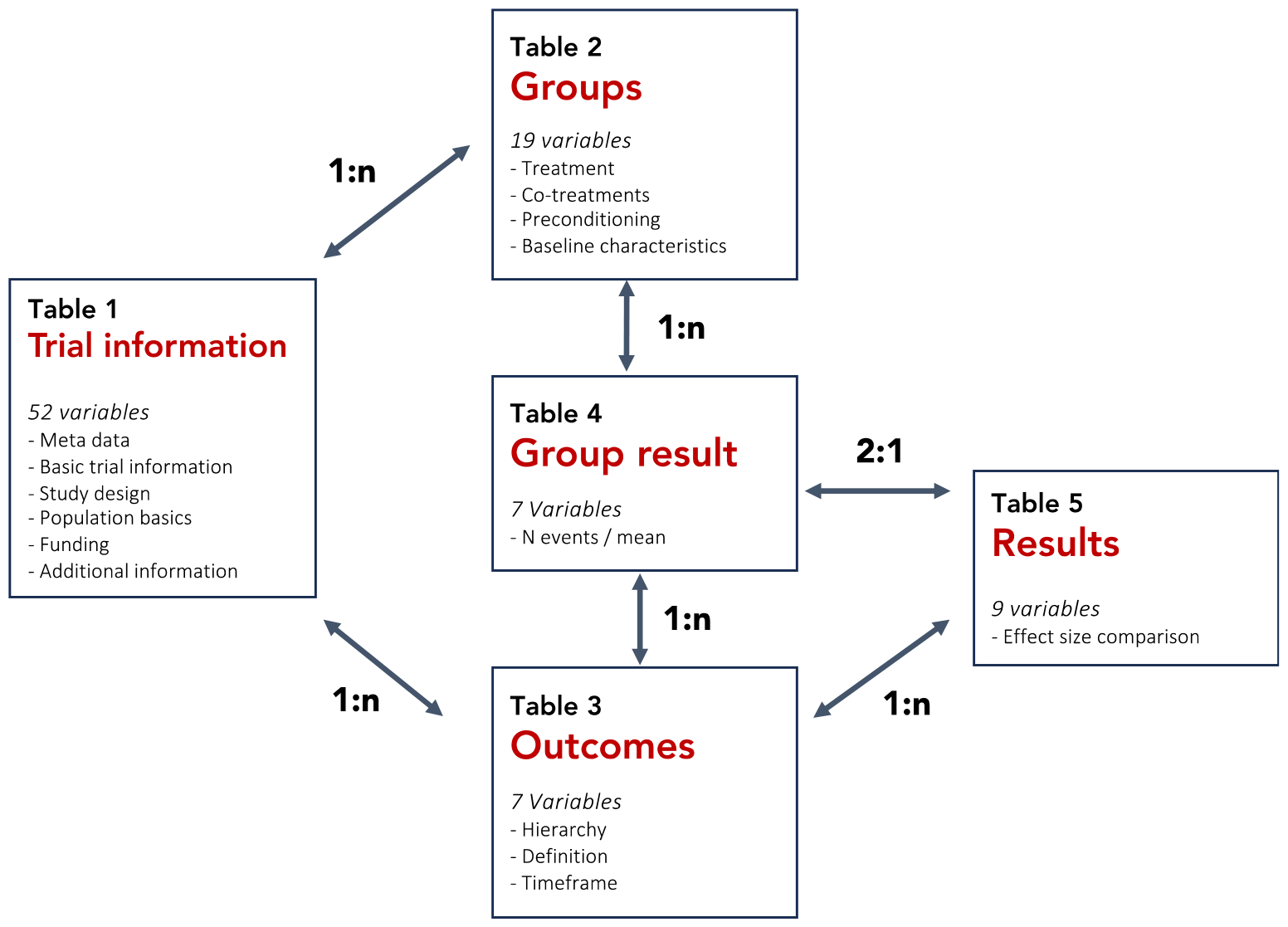
Overview of the relational database

We used the software application Directus (37) to access and curate the underlying database. Directus employs a user-friendly graphical interface, and the application is browser based, making it easy to use, especially in collaborative setups. The Directus interface allows to extract and curate data from all included trial records retrieved from both PubMed and AACT.

### Data entry and curation

The CIEL-Library contains both automatically and manually extracted data. Meta-data from PubMed retrieved publications, such as title, abstract, and year of publication were automatically imported. All other trial information (e.g. trial design, outcomes, and results) derived from PubMed retrieved publications require manual extraction by full-text assessment of the corresponding paper. Depending on the individual trial reporting, this may yield different level of information completeness in the CIEL-Library.

For ClinicalTrials.gov entry data, we were able to automatically import, for now, 46 variables from the AACT database. The main hindrance for automatic extraction is the available AACT data format. Some information on ClinicalTrials.gov is not coded in a format that allows unique identification and extraction through the AACT database. For example, when multiple interventions administered in a trial are reported in the same data field without defining which intervention is the main treatment, which is a co-treatment, and so forth.

Therefore, such information must be extracted manually subsequently to the automatic data import. Similarly, specific trial information in AACT (e.g. contact person or group description), are moved from one table to another depending on whether trial results are reported, which adds to the complexity, and confusion, of importing data effectively.

For the initial database implementation, we prioritized results extraction for the ‘objective response rate’ outcome (38). This is a commonly used efficacy outcome reported in interventional trials based on the Response Criteria in Solid Tumor (RECIST) framework (39). We made this choice for feasibility reasons to not overload the first manual data extraction phase; many different outcomes may be reported in cancer trial publications, and at multiple time points, which is time consuming to extract manually. Other efficacy outcomes and safety outcomes that are reported or specified on trial registries or publications were recorded in our database (in Table 3; outcomes) but the results were not systematically extracted and curated (in Table 4-5; group results and results).

### The CIEL-Library website and web app (workstream 4)

The CIEL-Library website, accessible at http://ciel-library.org, provides an overview of the project. The actual ***CIEL-Library*** (i.e. the library of clinical trials) and ***Match My Patient*** are available on the CIEL-Library web application, accessible at http://app.ciel-library.org/. The web application landing page contains a high-level dashboard showing information on the number of trials, patients, and interventions.

The ***CIEL-Library*** enables the user to search and filter all included immunotherapy trials based on six options (Figure 3): type of therapy (e.g. tumor-infiltration lymphocytes), trial status (e.g. ongoing or completed), trial phase (e.g. phase 1 or 2), study design (e.g. randomized or non-randomized), disease category (e.g. dermatology) and level of data curation (e.g. curated or automatically imported). The search results in a listing of relevant trials, with basic information for each trial visible in a glance including date of publication or registration, title, trial status (e.g. published or ongoing), link to full publication or registration entry, whether the trial is randomized or not, level of data curation and funding. Furthermore, information regarding the *Population* (i.e. the patient group), *Intervention* (i.e. the tested treatment), and *Comparator* (the reference treatment, if any) are also shown. By clicking on the trial, more granular baseline characteristics (i.e. age, sex, disease stage, pre-treatment, performance status, and location of the trial), intervention/comparator (i.e., concomitant and preconditioning treatments) and results information appear. We use color coding to indicate whether results are reported (green – reported; grey – not reported).

**Figure 3:**
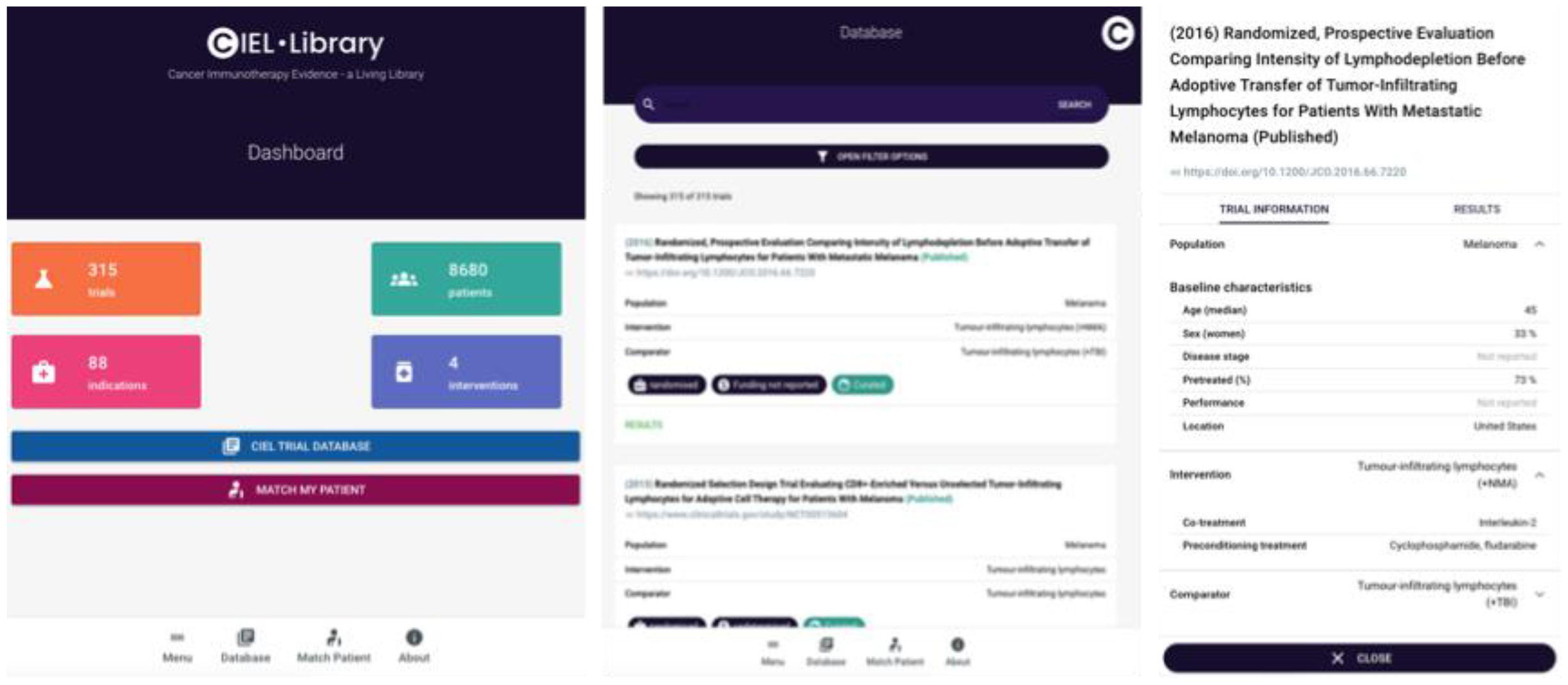
Overview of the CIEL-Library From left to right: landing page of the web App with a dashboard; view of the trials in the CIEL-Library with search bar and filters option; and a more granular view of the trial Population.

Currently, we show only some of the total variables collected to not overwhelm the user. The underlying database is available upon request allowing a more granular assessment of each trial for research purposes.

In ***Match My Patient***, _trials are identified through a three-step process (Figure 4). Step 1: Search for ongoing trials (i.e., to find a trial to enroll your patient) or for completed trials (i.e., to find results for making a therapeutic decision). Step 2: Select the patient’s main disease area, e.g. metastatic melanoma. Step 3: Specify patient characteristics; disease stage (stage 1 to 4; based on the TNM staging system (40)), pre-treatment status (treatment naïve; 1 to 3 previous cancer treatments, or 4+ cancer treatments), performance status (0 to 4, based on the Eastern Cooperative Oncology Group scale) (41), and finally the geographic location of the patient. The resulting relevant trials are then presented as described above. Any search criteria which match a relevant trial are colored in green.

**Figure 4:**
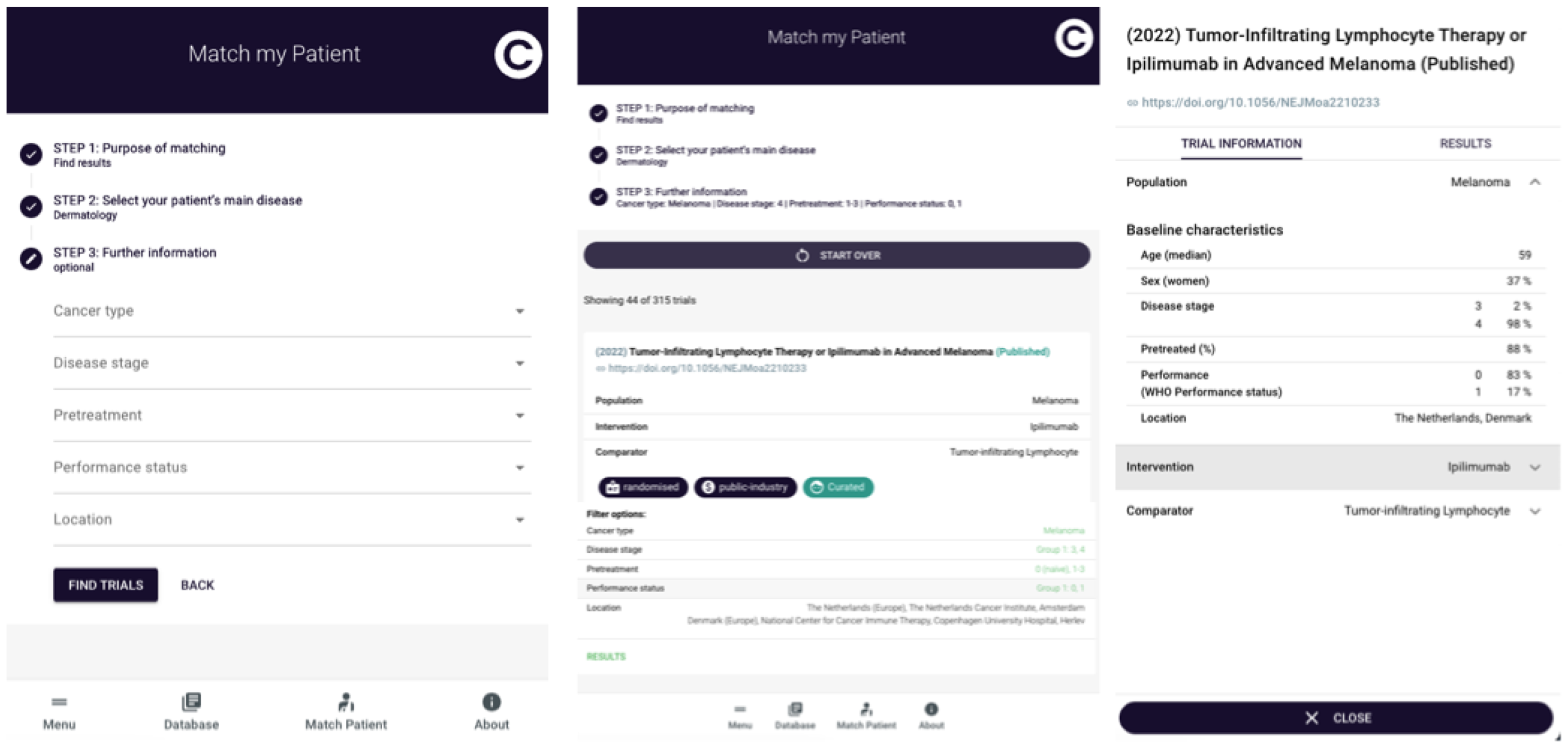
Overview of Match My Patient From left to right: Match My Patient search tool; relevant trials identified; the lower part shows the corresponding matching terms with the search; detail view of a relevant trials.

## 3. Discussion

CIEL’s semi-automated extraction process embedded in a relational database framework sets it apart from static evidence synthesis products, like regular systematic reviews, which may not be kept up-to-date or have narrowly defined scopes, e.g. one intervention for one type of disease.

Compared to known continuously updated clinical trial platforms (42–44), CIEL has integrated several semi-automated workstreams. The manual work required to extract data from publications and to ensure that it is constantly updated is resource demanding and will be challenging to sustain.

CIEL’s potential clinical application and its direct use of clinical trials as the ‘knowledge base’ has not, according to our knowledge, been described for other clinical decision support systems (18,19). Such systems have gained interest in recent years especially in complex specialties as oncology (45,46). CIEL’s *Match My Patient* mimics this concept, except that CIEL’s knowledge base consist of interventional clinical trials, which may be a more granular knowledge base, thus enabling, in theory, a more personalized treatment decision approach. The methodological (ecological) challenge when using CIEL is the matching of individual patient characteristics with aggregated trial results.

### CIEL’s next phases

In addition to continued reiterative improvements of the above outlined workstreams, the next phase should include a pilot testing of the CIEL-Library web application to collect user information and to validate (or refute) its usefulness as a clinical tool. Secondly, additional important data sources should be added, including the World Health Organization International Clinical Trial Registry Platform and the Embase database of published literature. Less critical, albeit potentially useful features, would be the implementation of a ranking algorithm for the trials identified using the *Match My Patient* tool and a dataset download feature to allow dataset reuse for further research. Beyond this, automatizing manual workflows, such as a full-scale integration of the PubMed API into ASReview to allow genuinely automated updates, will improve the database’s long-term sustainability.

### Limitations

Although the CIEL-Library is currently not a living library, all parts of the machinery have been thought an implemented to allow for a continuously updated database. However, the main challenge to CIEL’s viability and usefulness are the time and resources needed to manually extract, import, and curate data from heterogenous and incomplete data sources, both journal publications and trial registries. As stated already in 2001 (17), the prerequisite to designing a successful continuously updated trial database is access to “machine-readable” research literature. Twenty years later, the AACT initiative is the best, and only, attempt, at providing a structured clinical trial dataset, albeit limited to the data available on ClinicalTrials.gov. The emergence of large language models, such as ChatGPT, may open the possibilities of extracting otherwise unstructured text and data from journal publications into structured “machine-readable” formats (47,48).

Secondly, the CIEL-Library output can only be as good as the included data, which pertains both to reporting issues and to the methodological quality of the included trials (17)). Reporting of phase 1 cancer trials frequently deviate between trial registry and final publication (49), reporting of interventional phase 2 cancer trials in medical journals is in general poor (50), and harms reporting from phase 3 trials have been highlighted as being particularly poorly reported in journals (51) and deviating from trial registries and clinical study reports (52).

Journal publications are not reliable, exhaustive reports of trial information, and ideally, all information imported into CIEL should come from clinical trial registries. Due to the current publication paradigm, the lion’s share of trial data, especially pertaining to phase I and II trials, are made available in journal publications. Clinical trial registries, and the AACT database, enables a largescale automated data import. However, trial registry entries are not perfect records of trial information and results are often not reported (53,54). These issues are general issues of evidence-based healthcare and pertains not only to cancer trials.

Thirdly, the methodological quality of the included immunotherapy trials may limit CIEL’s usefulness. It is well-documented that pivotal cancer trials used for drug approvals have methodological limitations. These include the use of single-arm designs without a comparator; the use of inappropriate and inferior comparators, reporting of surrogate outcomes and not patient-reported quality of life and overall survival, and high risk of bias (55,56). The CIEL-Library will provide the data to investigate these issues in immunotherapy trials in research projects, but CIEL will not provide any critical literature appraisal itself, such as risk of bias assessments.

## Conclusion

The CIEL-Library offers a blueprint for future evidence synthesis. The current main limitations are the technical challenges related to aggregating heterogenous data sources, the time required to extract information from paper-based publications, and the general low reporting quality of the included trials.

The CIEL-Library infrastructure is applicable across fields, indications, and specialties. We hope that the medical community will gradually become less focused on paper-based publications as evidence-base and instead shift towards a more rigorous and complete trial registry reporting. Such an ideal reporting ecosystem would (theoretically) enable a fully automatable and machine-readable clinical trial dataset, which could be directly integrated into databases such as the CIEL-Library for the benefit of clinicians, policy makers, and patients.

## Data Availability

All data produced in the present study are available upon reasonable request to the authors

## Ethics approval

N/A

## Consent for publication

N/A

## Availability of data and materials

All information and data pertaining to the CIEL-Library can be found on the CIEL website (https://ciel-library.org).

## Competing interests

RC2NB (Research Center for Clinical Neuroimmunology and Neuroscience Basel) is supported by Foundation Clinical Neuroimmunology and Neuroscience Basel. RC2NB has a contract with Roche for a steering committee participation of LGH, unrelated to this work.

K.B., J.H., P.D., B.K., L.G.H. and P.J. declares no competing interest for this project.

H.L. received travel grants and consultant fees from BMS and Merck, Sharp and Dohme (MSD).

H.L. received research support from BMS, GlycoEra, and Palleon Pharmaceuticals. H.L. is a co-founder of Glycocalyx Therapeutics.

## Funding

The CIEL Project was funded the Basel Cancer League (KLbB-5577-02-2022).

## Appendix 1

1. PubMed search strategy

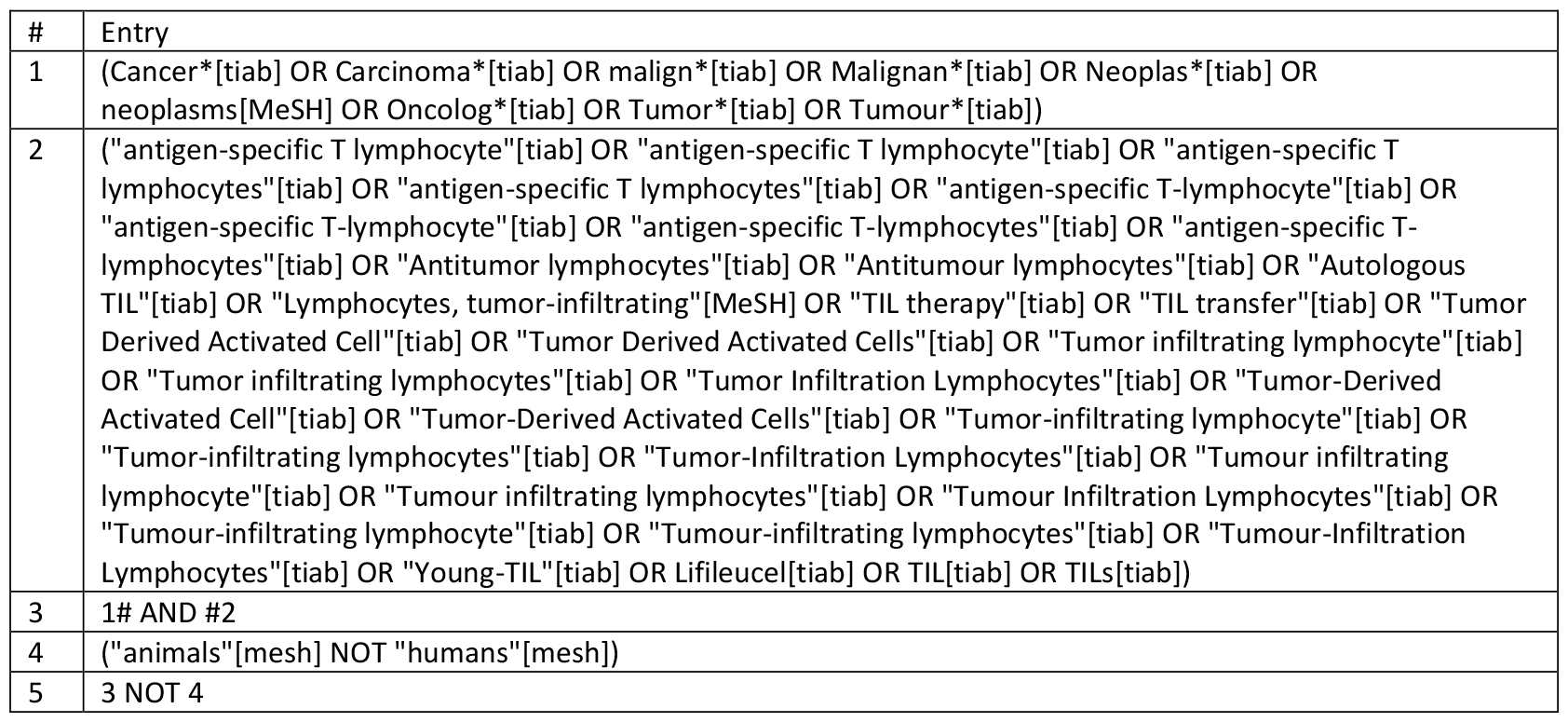

2. ClinicalTrials.gov search strategy; searched via Clinical Trials Transformation Initiative’s database (AACT) using the data tables ‘interventions.txt’ and ‘intervention_other_names.txt’ “antigen-specific T lymphocyte” OR “antigen-specific T lymphocytes” OR “antigen-specific T-lymphocyte” OR “antigen-specific T-lymphocytes” OR “Antitumor lymphocytes” OR “Antitumour lymphocytes” OR “Autologous TIL” OR “TIL therapy” OR “TIL transfer” OR “Tumor Derived Activated Cell” OR “Tumor Derived Activated Cells” OR “Tumor infiltrating lymphocyte” OR “Tumor infiltrating lymphocytes” OR “Tumor Infiltration Lymphocytes” OR “Tumor-Derived Activated Cell” OR “Tumor-Derived Activated Cells” OR “Tumor-infiltrating lymphocyte” OR “Tumor-infiltrating lymphocytes” OR “Tumor-Infiltration Lymphocytes” OR “Tumour infiltrating lymphocyte” OR “Tumour infiltrating lymphocytes” OR “Tumour Infiltration Lymphocytes” OR “Tumour-infiltrating lymphocyte” OR “Tumour-infiltrating lymphocytes” OR “Tumour-Infiltration Lymphocytes” OR “Young-TIL” OR Lifileucel OR TIL OR TILs

## References

1. Peri A, Salomon N, Wolf Y, Kreiter S, Diken M, Samuels Y. The landscape of T cell antigens for cancer immunotherapy. Nat Cancer. 2023 Jul;4(7):937–54.

2. Waldman AD, Fritz JM, Lenardo MJ. A guide to cancer immunotherapy: from T cell basic science to clinical practice. Nat Rev Immunol. 2020 Nov;20(11):651–68.

3. Rohaan MW, Wilgenhof S, Haanen JBAG. Adoptive cellular therapies: the current landscape. Virchows Arch Int J Pathol. 2019 Apr;474(4):449–61.

4. Farkona S, Diamandis EP, Blasutig IM. Cancer immunotherapy: the beginning of the end of cancer? BMC Med. 2016 May 5;14:73.

5. Cancer Research Institute [Internet]. [cited 2023 Dec 11]. Immuno-Oncology Landscape. Available from: https://www.cancerresearch.org/immuno-oncology-landscape

6. Saez-Ibanez AR, Upadhaya S, Campbell J. Immuno-oncology clinical trials take a turn beyond PD1/PDL1 inhibitors. Nat Rev Drug Discov. 2023 Jun;22(6):442–3.

7. Banerjee R, Prasad V. Characteristics of Registered Studies of Chimeric Antigen Receptor Therapies: A Systematic Review. JAMA Netw Open. 2021 Jul 1;4(7):e2115668.

8. Chen YJ, Abila B, Mostafa Kamel Y. CAR-T: What Is Next? Cancers. 2023 Jan 21;15(3):663.

9. Cancer Research Institute [Internet]. [cited 2023 Dec 11]. Cancer Vaccines. Available from: https://www.cancerresearch.org/treatment-types/cancer-vaccines

10. FDA Center for Biologics Evaluation and Research. IMLYGIC approval documents. FDA [Internet]. 2023 Feb 15 [cited 2023 Dec 11]; Available from: https://www.fda.gov/vaccines-blood-biologics/cellular-gene-therapy-products/imlygic

11. Straus SE, Sackett DL. Using research findings in clinical practice. BMJ. 1998 Aug 1;317(7154):339–42.

12. Bastian H, Glasziou P, Chalmers I. Seventy-five trials and eleven systematic reviews a day: how will we ever keep up? PLoS Med. 2010 Sep 21;7(9):e1000326.

13. Beller EM, Chen JKH, Wang ULH, Glasziou PP. Are systematic reviews up-to-date at the time of publication? Syst Rev. 2013 May 28;2:36.

14. Bashir R, Surian D, Dunn AG. Time-to-update of systematic reviews relative to the availability of new evidence. Syst Rev. 2018 Nov 17;7(1):195.

15. Khalil H, Ameen D, Zarnegar A. Tools to support the automation of systematic reviews: a scoping review. J Clin Epidemiol. 2022 Apr;144:22–42.

16. Marshall IJ, Wallace BC. Toward systematic review automation: a practical guide to using machine learning tools in research synthesis. Syst Rev. 2019 Jul 11;8(1):163.

17. Sim I, Gorman P, Greenes RA, Haynes RB, Kaplan B, Lehmann H, et al. Clinical decision support systems for the practice of evidence-based medicine. J Am Med Inform Assoc JAMIA. 2001;8(6):527–34.

18. Sutton RT, Pincock D, Baumgart DC, Sadowski DC, Fedorak RN, Kroeker KI. An overview of clinical decision support systems: benefits, risks, and strategies for success. NPJ Digit Med. 2020;3:17.

19. Kwan JL, Lo L, Ferguson J, Goldberg H, Diaz-Martinez JP, Tomlinson G, et al. Computerised clinical decision support systems and absolute improvements in care: meta-analysis of controlled clinical trials. BMJ. 2020 Sep 17;370:m3216.

20. COVID-evidence – Evidence [Internet]. [cited 2023 Dec 11]. Available from: https://covid-evidence.org/

21. PragMeta – Generalizability, applicability, and pragmatism of clinical trials and their impact on treatment effect estimates: a meta-epidemiological study [Internet]. [cited 2023 Dec 11]. Available from: https://pragmeta.org/

22. Hirt J, Janiaud P, Düblin P, Hemkens LG. Meta-research on pragmatism of randomized trials: rationale and design of the PragMeta database. Trials. 2023 Jun 30;24(1):437.

23. CEIT-cancer – The Comparative Effectiveness of Innovative Treatments for Cancer project [Internet]. [cited 2023 Dec 11]. Available from: https://ceit-cancer.org/

24. Ladanie A, Speich B, Naudet F, Agarwal A, Pereira TV, Sclafani F, et al. The Comparative Effectiveness of Innovative Treatments for Cancer (CEIT-Cancer) project: Rationale and design of the database and the collection of evidence available at approval of novel drugs. Trials. 2018 Sep 19;19(1):505.

25. Wang S, Sun J, Chen K, Ma P, Lei Q, Xing S, et al. Perspectives of tumor-infiltrating lymphocyte treatment in solid tumors. BMC Med. 2021 Jun 11;19(1):140.

26. Paijens ST, Vledder A, de Bruyn M, Nijman HW. Tumor-infiltrating lymphocytes in the immunotherapy era. Cell Mol Immunol. 2021 Apr;18(4):842–59.

27. Dafni U, Michielin O, Lluesma SM, Tsourti Z, Polydoropoulou V, Karlis D, et al. Efficacy of adoptive therapy with tumor-infiltrating lymphocytes and recombinant interleukin-2 in advanced cutaneous melanoma: a systematic review and meta-analysis. Ann Oncol Off J Eur Soc Med Oncol. 2019 Dec 1;30(12):1902–13.

28. Chen B, Li H, Liu C, Xiang X, Wang S, Wu A, et al. Prognostic value of the common tumour-infiltrating lymphocyte subtypes for patients with non-small cell lung cancer: A meta-analysis. PloS One. 2020;15(11):e0242173.

29. Cancer Research Institute [Internet]. [cited 2023 Dec 11]. Cancer Cell Therapy Landscape | Cancer Research Institute (CRI). Available from: https://www.cancerresearch.org/cancer-cell-therapy-landscape

30. PubMed PubReMiner: a tool for PubMed query building and literature mining [Internet]. [cited 2023 Dec 11]. Available from: https://hgserver2.amc.nl/cgi-bin/miner/miner2.cgi

31. van de Schoot R, de Bruin J, Schram R, Zahedi P, de Boer J, Weijdema F, et al. An open source machine learning framework for efficient and transparent systematic reviews. Nat Mach Intell. 2021 Feb;3(2):125–33.

32. ASReview [Internet]. [cited 2023 Dec 11]. ASReview - Active learning for Systematic Reviews. Available from: https://asreview.nl/

33. Tasneem A, Aberle L, Ananth H, Chakraborty S, Chiswell K, McCourt BJ, et al. The database for aggregate analysis of ClinicalTrials.gov (AACT) and subsequent regrouping by clinical specialty. PloS One. 2012;7(3):e33677.

34. AACT Database | Clinical Trials Transformation Initiative [Internet]. [cited 2023 Dec 11]. Available from: https://aact.ctti-clinicaltrials.org/

35. Dilling T. Artificial Intelligence Research: The Utility and Design of a Relational Database System. Adv Radiat Oncol [Internet]. 2020 Jul 13 [cited 2023 Dec 11];5(6). Available from: https://pubmed.ncbi.nlm.nih.gov/33305089/

36. Boesen K, Hirt J, Hemkens LG, Düblin P, Janiaud P. The Cancer Immunotherapy Evidence Living (CIEL) Library project [Internet]. OSF; 2023 [cited 2023 Dec 18]. Available from: https://osf.io/emyhj/

37. Directus: The Backend to Build Anything or Everything [Internet]. [cited 2023 Dec 11]. Available from: https://directus.io/

38. Aykan NF, Özatlı T. Objective response rate assessment in oncology: Current situation and future expectations. World J Clin Oncol. 2020 Feb 24;11(2):53–73.

39. Eisenhauer EA, Therasse P, Bogaerts J, Schwartz LH, Sargent D, Ford R, et al. New response evaluation criteria in solid tumours: revised RECIST guideline (version 1.1). Eur J Cancer Oxf Engl 1990. 2009 Jan;45(2):228–47.

40. Brierley JD, Gospodarowicz MK, Wittekind C. TNM Classification of Malignant Tumours, 8th Edition | Wiley [Internet]. [cited 2023 Dec 11]. Available from: https://www.wiley.com/en-es/TNM+Classification+of+Malignant+Tumours%2C+8th+Edition-p-9781119263579

41. Oken MM, Creech RH, Tormey DC, Horton J, Davis TE, McFadden ET, et al. Toxicity and response criteria of the Eastern Cooperative Oncology Group. Am J Clin Oncol. 1982 Dec;5(6):649–55.

42. WWARN Clinical Trials Publication Library | Infectious Diseases Data Observatory [Internet]. [cited 2023 Dec 11]. Available from: https://www.iddo.org/wwarn/wwarn-clinical-trials-publication-library

43. Evidence Map - The Map of Pancreatic Surgery [Internet]. [cited 2023 Dec 11]. Available from: https://www.evidencemap.surgery/

44. Evidence Finder - Orygen, Revolution in Mind [Internet]. [cited 2023 Dec 11]. Available from: https://orygen.org.au/Training/Evidence-Finder

45. Nafees A, Khan M, Chow R, Fazelzad R, Hope A, Liu G, et al. Evaluation of clinical decision support systems in oncology: An updated systematic review. Crit Rev Oncol Hematol. 2023 Dec;192:104143.

46. Pawloski PA, Brooks GA, Nielsen ME, Olson-Bullis BA. A Systematic Review of Clinical Decision Support Systems for Clinical Oncology Practice. J Natl Compr Cancer Netw JNCCN. 2019 Apr 1;17(4):331–8.

47. Roberts RH, Ali SR, Hutchings HA, Dobbs TD, Whitaker IS. Comparative study of ChatGPT and human evaluators on the assessment of medical literature according to recognised reporting standards. BMJ Health Care Inform. 2023 Oct;30(1):e100830.

48. Qureshi R, Shaughnessy D, Gill KAR, Robinson KA, Li T, Agai E. Are ChatGPT and large language models “the answer” to bringing us closer to systematic review automation? Syst Rev. 2023 Apr 29;12(1):72.

49. Shepshelovich D, Goldvaser H, Wang L, Abdul Razak AR, Bedard PL. Comparison of reporting phase I trial results in ClinicalTrials.gov and matched publications. Invest New Drugs. 2017 Dec;35(6):827–33.

50. Grellety T, Petit-Monéger A, Diallo A, Mathoulin-Pelissier S, Italiano A. Quality of reporting of phase II trials: a focus on highly ranked oncology journals. Ann Oncol Off J Eur Soc Med Oncol. 2014 Feb;25(2):536–41.

51. Sivendran S, Latif A, McBride RB, Stensland KD, Wisnivesky J, Haines L, et al. Adverse event reporting in cancer clinical trial publications. J Clin Oncol Off J Am Soc Clin Oncol. 2014 Jan 10;32(2):83–9.

52. Paludan-Müller AS, Créquit P, Boutron I. Reporting of harms in oncological clinical study reports submitted to the European Medicines Agency compared to trial registries and publications-a methodological review. BMC Med. 2021 Apr 8;19(1):88.

53. Goldacre B, DeVito NJ, Heneghan C, Irving F, Bacon S, Fleminger J, et al. Compliance with requirement to report results on the EU Clinical Trials Register: cohort study and web resource. BMJ. 2018 Sep 12;362:k3218.

54. DeVito NJ, Bacon S, Goldacre B. Compliance with legal requirement to report clinical trial results on ClinicalTrials.gov: a cohort study. Lancet Lond Engl. 2020 Feb 1;395(10221):361–9.

55. DeLoughery EP, Prasad V. The US Food and Drug Administration’s use of regular approval for cancer drugs based on single-arm studies: implications for subsequent evidence generation. Ann Oncol Off J Eur Soc Med Oncol. 2018 Mar 1;29(3):527–9.

56. Kim C, Prasad V. Strength of Validation for Surrogate End Points Used in the US Food and Drug Administration’s Approval of Oncology Drugs. Mayo Clin Proc. 2016 May 10;S0025-6196(16)00125-7.

